# Military participation in health security: analysis of Joint External Evaluation mission reports and National Action Plans for Health Security

**DOI:** 10.1101/2020.04.21.20073270

**Authors:** Brett M. Forshey, Alexandra K. Woodward, Jose L. Sanchez, Stephanie R. Petzing

**Author notes:** These authors contributed equally to this manuscript (Brett M. Forshey, Alexandra K. Woodward).

## Abstract

Militaries across the world play an important but at times poorly defined and underappreciated role in global health security. For example, they are often called upon to support civilian authorities in humanitarian crises and to provide routine healthcare for civilians. Furthermore, military personnel are a unique population in a health security context, as they are highly mobile and often deploy to austere settings domestically and internationally, which may increase exposure to infectious diseases. Despite the role of militaries, few studies have systematically evaluated the involvement of militaries in global health security activities, including the Global Health Security Agenda (GHSA). To address this shortcoming, we analyzed Joint External Evaluation (JEE) mission reports (n=91) and National Action Plans for Health Security (n=11) that had been completed as of October 2019 (n=91) to determine the extent to which military organizations have been involved in the evaluation process, country military contributions to health security are accounted for, and specific recommendations are provided for the country’s military. For JEE reports, military involvement was highest for the “Respond” core area (73%) but much lower for the Prevent (36%) and Detect (30%) core areas. Similarly, 73% of NAPHS documents mentioned military involvement in the Respond core area, compared to 27% and 36% for Prevent and Detect, respectively. Additionally, only 26% of JEE reports provide recommendations for the military in any of the core areas. Our results indicate the need to more fully incorporate military roles and contributions into the GHSA framework and other health security activities in order to improve national capabilities to prevent, detect, and respond to infectious disease threats.

## Introduction

In response to increasing risks of international spread of infectious diseases, countries around the world agreed to the revised International Health Regulations (2005) (IHR 2005), which came into force in 2007. The goal of the IHR 2005 was to protect against the international spread of diseases while avoiding undue restrictions on international traffic and trade. The IHR 2005 requires participating countries to strengthen their capabilities in 19 health-related technical areas, including disease surveillance and response capabilities. Despite the widespread agreement to participate, many countries have been unable to meet the goals established in the IHR 2005 (Gostin, Katz, 2016).

In 2014, the Global Health Security Agenda (GHSA) was introduced to re-invigorate action toward the IHR 2005 goals and establish partnerships to build country capacities. The GHSA is organized around 11 Action Packages to prevent, detect, and respond to public health emergencies, particularly infectious disease threats, with goals that line up with IHR 2005 objectives (see WHO, 2008; Katz, Sorrell, et al 2014.) The GHSA explicitly encourages a multisectoral approach to capacity building, with the goal of being able to conduct multisectoral response to natural or intentional biological events, linking public health and law enforcement, and developing and implementing a framework for sharing information across human and animal health, law enforcement, and the defense sector.

Global health security is relevant to the military establishment for multiple reasons, including the impact of infectious diseases on operational readiness and because of the support that militaries can provide to the civilian sector during outbreaks of infectious diseases and other public health emergencies, especially with regards to enhancement of public health surveillance and response capabilities. Infectious diseases have had, and continue to have, a profound impact on military operations. As an example, during World War II, malaria infections affected at least 124,000 U.S. military personnel in the Pacific (Ho, Hwang, and Lee 2014). More recently, HIV/AIDS reduced troop strength for countries across sub Saharan Africa (Feldbaum, Lee, and Patel 2006) In addition to these two large scale examples, regular outbreaks of malaria and other febrile illnesses, diarrheal diseases, and respiratory diseases have been known to limit operational readiness for militaries around the world (Zemke et a.l 2019; Chretien et al. 2007; Sebeny et al 2013).

Military personnel represent a unique population from a health security perspective. They are typically highly mobile, self-sustaining and often are able to deploy to austere settings, domestically and internationally, which may increase exposure to emerging pathogens. Recruits train in crowded conditions, increasing risk for pathogen transmission. Military personnel are also often immunologically naive to circulating pathogens (Institute of Medicine, 2002), which puts them at elevated risk for infections and diseases, such as malaria (Manning et al 2014). Not only are military personnel at risk for infections while deployed, these exposures can pose a risk for host nation populations due to transmission of infectious pathogens; similarly these pathogens may represent a risk for the military personnels’ own countries upon redeployment back to their home of origin (Zemke 2019). Examples abound where military personnel have been involved with disease transmission events during deployment internationally or during exercises within their national borders. Spread of pandemic influenza, both in 1918 and in 2009, was likely exacerbated by in-country and international travel of military personnel (Johns, Blazes et al. 2011). Notably, cholera was introduced into Haiti following the 2010 earthquake, most likely by Nepalese peacekeepers (Frerichs, Keim et al. 2012, Eppinger, Pearson et al. 2014, Orata, Keim et al. 2014), leading to a large outbreak (hundreds of thousands of cases) of diarrheal disease among a highly susceptible host nation civilian population. Mobility can be a concern even within a country’s boundaries. For example, an outbreak of malaria in the northern coast of Peru, in a region where no malaria had been reported for four years, was linked to *Plasmodium falciparum* strains potentially introduced by military personnel returning from operations in the Peruvian Amazon (Baldeviano, Okoth et al. 2015).

Militaries have a clear responsibility to provide public health support and health care to their troops; however, they also often provide support to the civilian sector in many ways. Many military medical clinics and hospitals provide extensive medical treatment to their countries’ civilian populations, a relationship that is often overlooked in country-level health security initiatives. In many countries, “the military has the strongest medical infrastructure and capability that should be leveraged.” (Russell, Johns et al. 2011) Among African militaries surveyed, as much as 50% to 80% of delivered healthcare by military medical services is provided to civilians with no military affiliation (McCollum, Hanna et al. 2015) Similarly, the Jordanian Military Royal Medical Services runs 10 hospitals that serve approximately 30% of their civilian population (Al-Qudah 2011, World Health Organization and Ministry of Health - Jordan 2011). Militaries provide support to civil authorities internally and internationally when civilian capabilities are exceeded, particularly in humanitarian relief or times of conflict, as occurred during the 2014-2015 West Africa Ebola outbreak (Kamradt-Scott 2016). Militaries can provide a range of capabilities in security, transportation, logistics, and communications (Licina 2011; Michaud, Moss, et al. 2019), and, less frequently, direct patient care, as happened during the Middle East respiratory syndrome coronavirus (MERS-CoV) outbreak in the Republic of Korea. The military often also plays a critical role in national biodefense issues, due to the need to prevent or respond to the potential misuse of biological agents as bioweapons (Fidler 2011).

To examine the incorporation of each GHSA member country’s military into their GHSA activities, we examined two types of reports that aim to assess and build country capacity for public health surveillance and response to meet IHR 2005 requirements. First, we analyzed the World Health Organization’s (WHO) Joint External Evaluations (JEEs). Through the JEE process, experts work with each country to assess the country’s abilities to prevent, detect, and respond to all-hazards public health threats, including infectious diseases, and provide recommendations for improvement (Bell, Tappero et al. 2017). Second, we evaluated available National Action Plans for Health Security (NAPHS), which outline country priorities for addressing shortcomings identified in the JEEs and improving public health capabilities.

## Methods

We systematically screened and analyzed the available JEE reports and NAPHS for mentions of country military involvement. Two analysts evaluated all reports, with both analysts looking at each report to verify the data collected. Any discrepancies between the two analysis were resolved by further review of the document in question. Reports were screened for mentions of military involvement, in part through keyword searches for the following terms (or their equivalents in French): “military,” “defens(c)e”, “army,” “navy,” “air force,” “MoD,” “armed forces,” as well as any acronyms identified in the beginning of each report that represented the country’s military. Militaries were defined similarly to Michaud et al (Michaud, Moss et al. 2019): organized armed groups tasked with providing defense and prosecuting war on behalf of the respective countries. Civil defense organizations and other domestic security agencies distinct from the country’s military were not included.

### JEE Report Analysis

All JEE mission reports from the six WHO regions that had been published online (http://www.who.int/ihr/procedures/mission-reports/en/) as of October 2019 (n=91) were analyzed; 76 were available in English and 15 were available only in French.

Reports were analyzed, in part using the keyword terms described above, based the following metrics:

- The report listed military participants;
- The total number of military participants (if listed in the report);
- The report mentioned specific examples of military-civilian engagement;
- The report referenced the current role of the military in each of the JEE core areas (Prevent, Detect, Respond, Other IHR-related hazards and Points of Entry or in the introduction);
- The report provided recommendations for future military engagement in each of the core areas;
- The report referenced any military policy documents related to global health security;
- The report mentioned collaboration of militaries from other countries, including any regional cooperation, and
- The evaluation team included any experts with listed military or Ministry of Defense affiliation

Once all reports were analyzed and reviewers came to concordant responses, each metric was evaluated across all reports and across reports within each WHO region - specifically Africa (AFRO), Eastern Mediterranean (EMRO), Europe (EURO), the Americas (PAHO), South East Asia (SEARO), and the Western Pacific (WPRO) - to determine proportions for each metric.

### NAPHS Reports

All NAPHS reports from the six WHO regions that had been published online (https://extranet.who.int/sph/country-planning) as of October 2019 (n=11) were evaluated (using the same keyword search methodology used to evaluate the JEE reports) to determine whether military involvement was mentioned in any of the JEE core areas: Prevent, Detect, Respond, and Other IHR-related hazards and Points of Entry (“Other”). Involvement of militaries from other countries was also examined in each report.

## Results

### JEE Reports

To conduct this analysis, we analyzed a total of 91 JEE reports (Table). The reports were divided between six WHO regions, with AFRO publishing the largest number of reports (n = 41), followed by EMRO (n = 17), EURO (n = 13), WPRO (n = 10), SEARO (n = 8), and PAHO (n = 2). Among the 91 reports across all WHO regions, slightly over half (n = 47, 52%) included military participation in the assessment process. Not all of these 47 reports included a list of participants; among the 24 that did include a list, the median number of military participants was one per report, with a range of one to 10 (Thailand) military personnel interviewees.

**Table.** References to military involvement in JEEs, by WHO region.

| **Table. References to military involvement in JEEs, by WHO region.** |  |  |  |  |  |  |  |
| --- | --- | --- | --- | --- | --- | --- | --- |
|  | **Percentage of countries (n)** |  |  |  |  |  |  |
| **References to military participation** | **Total**<br>**(n = 91)** | **AFRO**<br>**(n = 41)** | **EMRO**<br>**(n = 17)** | **EURO**<br>**(n = 13)** | **PAHO**<br>**(n = 2)** | **SEARO**<br>**(n = 8)** | **WPRO**<br>**(n = 10)** |
| Military participation in JEE process* | 57% (47) | 49% (18) | 67% (10) | 25% (3) | 100% (2) | 86% (6) | 80% (8) |
| Mil-civ engagement mentioned | 78% (71) | 71% (29) | 94% (16) | 77% (10) | 100% (2) | 75% (6) | 80% (8) |
| Role of mil/MoD referenced in at least one core area | 86% (78) | 85% (35) | 87% (13) | 92% (12) | 100% (2) | 75% (6) | 80% (8) |
| Prevent core area | 36% (33) | 27% (11) | 47% (8) | 62% (8) | 100% (2) | 25% (2) | 20% (2) |
| Detect core area | 30% (27) | 22% (9) | 59% (10) | 8% (1) | 100% (2) | 38% (3) | 20% (2) |
| Respond core area | 73% (66) | 76% (31) | 71% (12) | 77% (10) | 50% (1) | 75% (6) | 60% (6) |
| Other section or Introduction | 41% (37) | 29% (12) | 47% (8) | 54% (7) | 100% (2) | 38% (3) | 50% (5) |
| Recommendations for Mil/MoD provided in at least one core area | 26% (24) | 29% (12) | 13% (2) | 23% (3) | 0% (0) | 63% (5) | 20% (2) |
| Prevent core area | 8% (7) | 7% (3) | 6% (1) | 0% (0) | 0% (0) | 25% (2) | 10% (1) |
| Detect core area | 4% (4) | 0% (0) | 6% (1) | 8% (1) | 0% (0) | 0% (0) | 20% (2) |
| Respond core area | 16% (15) | 22% (9) | 6% (1) | 8% (1) | 0% (0) | 38% (3) | 10% (1) |
| Other section or Introduction | 5% (5) | 5% (2) | 6% (1) | 8% (1) | 0% (0) | 0% (0) | 10% (1) |
| Reference to relevant mil/MoD policy documents | 34% (31) | 37% (15) | 24% (4) | 46% (6) | 50% (1) | 13% (1) | 40% (4) |
| Any mention of involvement of militaries from other countries, including regional cooperation | 9% (8) | 2% (1) | 18% (3) | 8% (1) | 0% (0) | 13% (1) | 20% (2) |
| JEE team included individuals affiliated with a military organization | 7% (6) | 12% (5) | 0% (0) | 0% (0) | 50% (1) | 0% (0) | 0% (0) |

\* 8 of 91 did not list participating organizations

The majority of reports (n = 71, 78%) mentioned some level of military-civilian engagement in at least one section and mentioned the country’s military in at least one of the Prevent, Detect, Respond, and Other sections (n = 78, 86%). Across all reports, military engagement was most commonly mentioned in the Respond core area (n = 66, 73%), followed by the Other core area or Introduction (n = 37, 41%), the Prevent core area (n = 33, 36%), and the Detect core area (n = 27, 30%). Most military mentions were only cursory or included as part of list of national institutions (e.g. for Moldova: “…more than ten national authorities/institutions are involved in biosafety and biosecurity, including the Ministry of Defence”; https://www.who.int/ihr/publications/WHO-WHE-CPI-2019-54/en/).

Recommendations for enhanced military engagement in any of the core areas were infrequently made across all reports: only 24 (26%) mentioned enhancing military collaboration in any of the core area recommendation sections. Recommendations were most commonly seen in the Respond core area (n = 15, 16%) and least commonly seen in the Detect core area (n = 4, 4%). Only 31 of the reports (34%) referenced any policy documents related to the country’s military. Eight reports (9%) referenced the involvement of militaries from other countries (which included the mention of funding support to conduct activities in the core areas as well as joint exercises and outbreak collaboration), and only six reports (7%) included individuals on the JEE assessment team who held positions specifically affiliated with military or defense organizations.

### Regional and Cross-Regional Findings

We explored the metrics used in our analysis from a regional and cross-regional perspective (Table). Military participants were included in the majority of country assessments in the WPRO (80%), SEARO (75%), and EMRO (59%) regions, and were less frequently included in country assessments in the AFRO (44%) and EURO (23%) regions. Consistent with the overarching findings across all regions, military-civilian engagement was mentioned in the majority of reports in each region, with the greatest mention of military-civilian engagement identified for the EMRO region (94%) relative to the AFRO (71%) and SEARO regions (75%).

The role of the military was mentioned for at least one of the core areas in over 80% of all country assessments in each region, except SEARO (75%). Also consistent with overarching findings across all regions, military involvement was most frequently mentioned in the Respond core area in most regions. Recommendations for enhanced military engagement were infrequently made in country assessments in each region, except SEARO (5/8, 63%). The highest proportion of reports including at least one mention of any military-related policy documents was seen in the EURO reports (46%).

### NAPHS Reports

Reports from 11 countries (Australia, Benin, Eritrea, Liberia, Myanmar, Nigeria, Sierra Leone, Sri Lanka, Tanzania, Uganda, and the United States) were available for evaluation. Overall, 73% (8/11) listed some level of military participation in the NAPHS development process. Plans for military involvement were included for 27% (3/11) for Prevent, 36% (4/11) for Detect, 73% (8/11) for Respond, and 45% (5/11) for other IHR-related hazards. Of the countries with NAPHS and JEE reports for evaluation, military involvement was mentioned in both documents for 9% (1/11) for Prevent, 9% (1/11) for Detect, 64% (7/11) for Respond, and 18% (2/11) for Other core areas.

## Discussion

While militaries’ contributions were mentioned in at least one core area for the majority of country JEEs (86%), most mentions were brief and limited in scope. Far fewer JEEs (26%) provided recommendations for further military involvement in at least one of the core areas, most of which (63% of recommendations for militaries) were in the Respond core area. While the GHSA properly calls to link public health with law and multisectoral rapid response, including defense and other stakeholders within the security sector, there is insufficient incorporation of military involvement more broadly into Prevent and Detect core areas, which address priorities such as monitoring antimicrobial resistance, immunization programs, infectious disease surveillance, and public health workforce development. Further, comments and recommendations related to multisectoral engagement may be intended to include the military but are not clearly stated, which may limit the ability of the country to respond to JEE recommendations in a meaningful way. In fact, the limited military incorporation in the JEEs was also apparent in the NAPHSs, which are intended to drive country global health security activities over multiple years (WHO 2018).

### Recommendations

Our findings support several recommendations that are specific to GHSA engagement and speak more broadly to military engagement in global health security. We found that military involvement in Prevent and Detect core areas was quite limited. Military organization participation should extend beyond Respond core component Action Packages and should encompass all aspects of the Prevent and Detect Action Packages. For example,vaccination and prophylaxis plans, guidance, and requirements could include MoD-generated policies for their military personnel, and surveillance for infectious diseases among military personnel prior to and during deployments could be a point of emphasis (Chretien, Blazes, et. al 2007, Page; Faulde et al. 2010, de Laval, Dia et al. 2013; Sebeny and Chretien 2013). Additionally, since military health systems often support civilian populations, where applicable, military staff should be included in the workforce development aspects of GHSA to have the capacity to respond properly (Fidler 2011).

The Respond core area contained the most mention of military involvement. However, descriptions of military involvement were often vague and included recommendations for the military specifically for few (16%) countries. Future planning around the security sector should be specific about military involvement in future GHSA activities, which might include support for logistic, medical, and transportation capabilities related to preparedness, emergency response, and medical countermeasure deployment, where appropriate (McPhee et al 2019). We also found that few JEEs listed policy documents that explicitly reference the military, which indicates that military-civilian engagement may need to be clearly codified within government policies. Additionally, military response plans could be integrated into civilian government agencies plans and joint exercises could be established to ensure effective collaboration (Michaud, Moss et al. 2019). While initial points of within-country military-civilian engagement may revolve around biodefense issues and risk reduction of a particular disease or pathogen, these initial engagements could serve as springboards and building blocks for broader military-civilian health security engagement.

We found limited involvement by military personnel in the JEE process, particularly on the visiting JEE teams. Military authorities should be involved as experts on host country and external reviewer panels in future GHSA activities. Additionally, military-military engagements between GHSA partner countries should be considered to advance capabilities, since military-military partnerships may be more politically feasible in many instances. As an example of such activities, U.S. Department of Defense (DoD) has been actively engaged with partner militaries on health security activities aimed at countering biological threats and improving surveillance data for force health protection (Morrison, Ayotte, and Gerberding 2019; Quinnan 2016). These collaborations enable partner militaries to enhance their own capabilities to mitigate threats (Chretien, Blazes, et al. 2007). In Africa, an example of such collaboration is the African Partner Outbreak Response Alliance, where partner militaries have increased coordination to improve national and regional responses to infectious diseases. In Southeast Asia, malaria has been called out as a particular opportunity and need for military-military coordination, since militaries operate in remaining endemic regions and could collaborate to support malaria elimination efforts in often remote settings (Manning, Satharath et al. 2014). To ensure efficacy of these military-military engagements, metrics should be developed to monitor and evaluate military involvement in global health security (Ijaz, Kasowski et al. 2012; DoD Instruction 2000.30), since by and large, effects of military investment in global health have not been quantified (Licina 2011).

### Limitations

This study is not without limitations. The analysis is dependent on the content of the JEEs and NAPHS reports. In the case of the JEEs, the descriptions provided by the external reviewers may not reflect the full scope of participation of the country’s military. Additionally, the JEEs are subjective and largely qualitative (Ravi et al BMC Public Health) and are not designed for comparisons across countries and regions. However, the JEEs reflect the priorities of the JEE and country team, which shows that militaries were not a major focus. This may have been due to the lack of representatives from the defense organizations on the JEE panels or a lack of understanding of the role the defense sector can play in health security. Further, other studies have demonstrated the validity of the JEEs. For example, JEE scores are generally representative of other health outcomes and align with measures associated with public health system capabilities (Samhouri, Ijaz et al. 2018; Gupta, Kraemer et al. 2018), and JEEs corresponded with response capabilities during an outbreak (Garfield, Bartee et al. 2019). Additionally, JEEs provide a different perspective from self-evaluation (Tsai and Katz 2018; Tsia and Turba 2020), and our results from the JEEs is largely confirmed in the country plans laid out in the NAPHS.

Another limitation is that we utilized policy document titles mentioned in the JEEs and NAPHSs to determine if the documents referenced military involvement rather than reviewing each policy reference in detail. A deeper dive into the referenced policy documents may shed light on country policies that guide military engagement.

### Challenges to Involvement of Country Military Organizations

Involving military organizations in global health security activities is not without challenges, including political concerns, operational security, and funding. In the public health realm, the military establishment will always struggle to engage with the broader public health community because of the reality that the primary mission of militaries is not global health. While this is clearly a legitimate concern, militaries bear responsibility for the health of their troops and oftentimes the health of civilian populations. Additionally, the perceived militarization and “oversecuritization” of public health activities could cause alarm with the public and other stakeholders, including non-governmental organizations, thereby reducing buy-in, creating resistance among the public, and jeopardizing long-term execution of global health security activities (Wenham 2019). Cultural differences between health and military and policy restrictions may also complicate communication and limit military participation in civil affairs in many countries. GHSA helps to address these issues by creating a platform for collaboration across sectors, under civilian leadership using shared language and common goals.

National sovereignty may also be an issue when a military operating in a foreign country experiences or detects an infectious disease outbreak. For example, during the influenza A(H1N1) pandemic, the United States deferred to host countries regarding official reporting of influenza A(H1N1) cases among U.S. military personnel. However, based on IHR 2005, the U.S. would be obligated to report if the host country did not (Johns, Blazes et al. 2011). Clearly, military forces operating as peacekeepers or during exercises need to collaborate with the host country in reporting infectious disease cases, and be aware of potential for carrying pathogens back to their home country (Zemke 2019; Guerra, Ore, *et al*. 2019).

Operational security poses another challenge. During drafting of the IHR 2005, the U.S. agreed to/ratified the regulation to notify the WHO of public health risks occurring outside of its territory that may result in the international spread of disease, with the caveat that “any notification that would undermine the ability of the U.S. Armed Forces to operate effectively in pursuit of U.S. national security interests would not be considered practical for purposes of this Article” (WHO 2008). Notably, the Republic of Iran objected to this caveat, appealing to the universality of the treaty and stating that there is “no room for exempting the American Armed Forces, in particular those operating abroad.” Further, Iran claimed that the majority of WHO member states rejected the U.S.’s caveat. This concern about operational security, and controversy about the topic, no doubt applies to GHSA-related activities. Even WHO member states that disagreed with the caveat in IHR 2005 should be considered unlikely to fully comply with the notification requirements if operational security might be jeopardized.

Funding represents a particular challenge for supporting military participation in global health security capacity building. Funding to militaries may be perceived as military aid and may have legal restrictions at the federal level (Manning, Satharath et al. 2014). For example, under the President’s Malaria Initiative, and with USAID funding in general, no direct funds can be transferred directly to militaries (U.S. Government Accounting Office 2001); instead, the Ministry of Defense must participate in the Ministry of Health-led National Malaria Control Plan. Therefore, military participation is dependent on cooperation with and funds made available from often resource-restricted health ministries, which may serve as a barrier to access. Additionally, multinational organizations and non-governmental organizations may shy away from providing funding to support military public health activities.

## Conclusions

Despite the challenges and concerns about the role of the defense sector in global health security, it is important to recognize that militaries throughout the world are already engaged in these activities, with increasing involvement both within national borders and internationally (Wenham 2019; Michaud et al 2019). As emerging infectious disease threats are increasingly considered a national security and economic development concern (Fidler 2011), the role of militaries will likely continue to increase. Therefore, the question becomes how to constructively incorporate their military’s capabilities into each country’s framework to improve whole-of-country capability, while remaining consistent with national priorities and international guidelines (Michaud et al 2019). Additionally, military personnel are part of an increasingly connected world - United Nations peacekeeping operations alone involve more than 85,000 uniformed personnel from 122 countries participating in missions in diverse countries such as Haiti, Mali, and Kosovo (United Nations 2019) - and can be a potential source of infectious disease transmission across international boundaries (Zemke et al 2019). During future GHSA activities, military roles and contributions need to be included in all aspects of global health security and whole-of-government approaches to Prevent, Detect, and Respond to infectious diseases and other public health threats.

## Data Availability

All data used in this manuscript were extracted from publicly available reports from the WHO:
http://www.who.int/ihr/procedures/mission-reports/en/
https://extranet.who.int/sph/country-planning

## Disclaimer

The views expressed in this article are those of the authors and do not reflect the official policy or position of the Department of Defense or of the U.S. Government.

